# A Convolutional Neural Network based system for classifying malignant and benign skin lesions using mobile-device images

**DOI:** 10.1101/2023.12.06.23299413

**Authors:** Rim Mhedbi, Hannah O. Chan, Peter Credico, Rakesh Joshi, Joshua N. Wong, Collin Hong

**Affiliations:** York University, Toronto, Canada; Skinopathy Research, Skinopathy Inc., North York, Ontario, Canada; Scarborough Health Network, Centenary Hospital, Scarborough, Ontario, Canada; University of Alberta Hospital, Edmonton, Alberta, Canada

**Keywords:** Skin lesions, CNN, Error analysis, Melanoma, classification, Efficient Net, Camera images

## Abstract

The escalating incidence of skin lesions, coupled with a scarcity of dermatologists and the intricate nature of diagnostic procedures, has resulted in prolonged waiting periods. Consequently, morbidity and mortality rates stemming from untreated cancerous skin lesions have witnessed an upward trend. To address this issue, we propose a skin lesion classification model that leverages the efficient net B7 Convolutional Neural Network (CNN) architecture, enabling early screening of skin lesions based on camera images. The model is trained on a diverse dataset encompassing eight distinct skin lesion classes: Basal Cell Carcinoma (BCC), Squamous Cell Carcinoma (SCC), Melanoma (MEL), Dysplastic Nevi (DN), Benign Keratosis-Like lesions (BKL), Melanocytic Nevi (NV), and an ‘Other’ class. Through multiple iterations of data preprocessing, as well as comprehensive error analysis, the model achieves a remarkable accuracy rate of 87%.

## I. Introduction

Skin cancer is the most common type of cancer globally, with 1.5 million reported cases in 2020. Roughly 1/5 of these cases are melanomas [1]. Melanoma is the most lethal skin cancer with a mortality rate of 95%. However, if detected and treated early mortality rate drops to 5% [2]. Therefore, a lot of attention in the medical field has shifted toward the early detection and diagnosis of skin cancer. Various techniques have been adopted including the visual and manual inspection of the skin lesion using various methodologies, such as using the ABCDE lesion characterization rule when inspecting dermoscopic images [3]. Dermoscopic images are photographed using magnification and lighting using a dermatoscope, which can capture detailed skin structures in the epidermis that are not visible to the naked eye. Biopsy of the lesion is still the gold standard for verification of lesion identity. However, the aforementioned techniques require dermatology expertise, a dermatoscope, is time-consuming, and is still prone to error.

Lately, there has been a concerted effort to digitize skin lesion detection. With the rise of computer vision technology, many machine learning and deep learning algorithms were devised to systematically and automatically detect and diagnose skin lesions from dermoscopic images. [4] provided a survey of the most used techniques for skin lesion segmentation and classification from dermoscopic images. It was reported that CNN ensembles provided the best results with an accuracy ranging between 95% and 97% on The International Skin Imaging Collaboration (ISIC) dataset (a dataset that contains labeled dermoscopic images of different skin conditions). Further, it was reported that segmening images before classifying them resulted in major performance improvement. Similarly, using the ISIC dataset, [5] conducted a comparative study of various machine learning and deep learning models for skin lesion image classification. It was found that deep learning models significantly outperformed the conventional machine learning models (Xception net provided the highest performance with 86.45% accuracy).

Many other studies used the HAM10000 (“Human Against Machine with 10000 training images”) dataset which contains dermoscopic images of pigmented skin lesions. [6] proposed different approaches for skin lesion segmentation and classification. The skin lesions segmentation technique consists of using filtering and color transformation methods. The proposed skin disease classification procedure consists of applying two CNNs for feature extraction which are then fed to an SVM mode. Similarly, [7] proposed an architecture for skin lesion classification based on the HAM10000 dermoscopic images. Transfer learning was applied by using a pretrained DenseNet model for a skin lesion segmentation task. An ensemble of models was then created based on different versions of randomly balanced datasets obtained from the HAM10000. The proposed ensemble model achieved an accuracy of 0.899 and 0.785 in the validation set and test set, respectively.

CNNs therefore have displayed high performance in diagnosing skin lesions in dermoscopic images. However, as dermoscopic images are taken by dermatologists, making a doctor’s appointment is a prerequisite. Lately, due to a shortage of dermatologists and increased incidence of skin lesions, long wait times are common. This in turn can lead to delayed diagnosis and detrimental health repercussions. Camera images on the other hand can be easily available through the use of mobile devices. Diagnosing skin lesions based on camera images will help provide timely and early detection of skin lesions. However, based on the literature review conducted, we found that studies on Camera-based skin lesion classification were limited.

[3] proposed a framework for NMSC (Non-Melanoma Skin Cancer) detection based on camera images. First, the Elliptic Fourier Analysis (EFA) coefficients were obtained based on the skin lesion boundary. EFA coefficients along with asymmetry calculations characterized the irregularities in the shape of NMSC skin lesions. The computed coefficients acted as inputs into an SVM model with a Radial Basis Function (RBF) kernel. It was reported that the model obtained an accuracy of 78%.

In contrast, [8] proposed a model for conjunctival melanoma detection using ocular surface images, i.e. detection of melanoma located in the eye. The dataset contained 398 camera images belonging to four classes, including melanoma and other benign ocular surface lesions. Two classification scenarios were considered, the first corresponded to binary classification while the second corresponded to multi-class classification. A GAN was used for data augmentation and several CNN models were trained. It was found that GAN data augmentation plus MobileNet gave the best performance with an accuracy of 87.5% for the four-class classification and 97.2% for the binary classification.

In short, classifying skin lesions based on camera images has limited exploration. More importantly, the available skin lesion classification models’ performance need further improvement to reach clinical relevance. In this study,we propose a CNN for skin lesions multi-class classification based on camera images.

This manuscript is structured as follows. Section two presents the methodology including the description of the dataset, the adopted CNN model architecture, the evaluation metrics, and an ad hoc testing description. Section three demonstrates the results and discussion and section four provides the conclusions and future works.

## II. Methodology

### A. Datasets and preprocessing

To conduct this research, we used two datasets. The first dataset contains skin lesion images collected from Skinopathy and collaborators in the University of Alberta/Alberta Health System, under research ethics boards from the University of Alberta and NRC-IRAP. This dataset comprises 1993 skin lesion camera images pertaining to 33 skin conditions. The ground truth of these images was confirmed by pathology reports. The second dataset was obtained from a private dermatology clinic in Denmark and consists of various cutaneous lesions’ camera images. This dataset contains 3000 images that cover more than 91 classes of skin pathologies. The image labels for this second dataset were assigned based on the clinicians’ expert opinions [9].

In our study, we focused on classifying benign and cancerous skin lesions that cover seven classes. The cancerous skin lesions include Basal Cell Carcinoma (BCC) which is a type of cancer that originates in the basal cell, Squamous Cell Carcinoma (SCC) which is a skin cancer targeting the squamous cells, and Melanoma (MEL) which develops in the melanocytes. It is important to reiterate that MEL is the most lethal skin cancer if left untreated [2]. Another pre-malignant skin lesion included was Dysplastic Nevi (DN) which are atypical nevi (moles). It is important to note that DN have a high risk of turning into melanoma [10]. The other benign skin lesions comprise Benign Keratosis-Like lesions (BKL) which include Solar Lentigines, Seborrheic Keratoses, and Actinic Keratosis. Melanocytic Nevi (NV) and Vascular (VASC) correspond to the two other benign skin lesions. The NV class covers common benign moles including Spitz Nevus, Compound Nevus, Intrademal Nevus, Blue nevus, Junctional Nevus, Intradermal w/cogenita, and Lentiginous compound. The VASC lesions correspond to two types, namely, Hemangiomas, and Pyogenic Granulomas. The latter skin conditions are related to abnormalities in skin vessels. We have as well added an “Other” class which includes skin lesions and other dermatological issues that don’t belong to any of the previous classes or to normal skin. Figure 1 presents example images of the aforementioned skin lesions.

**Fig. 1:**
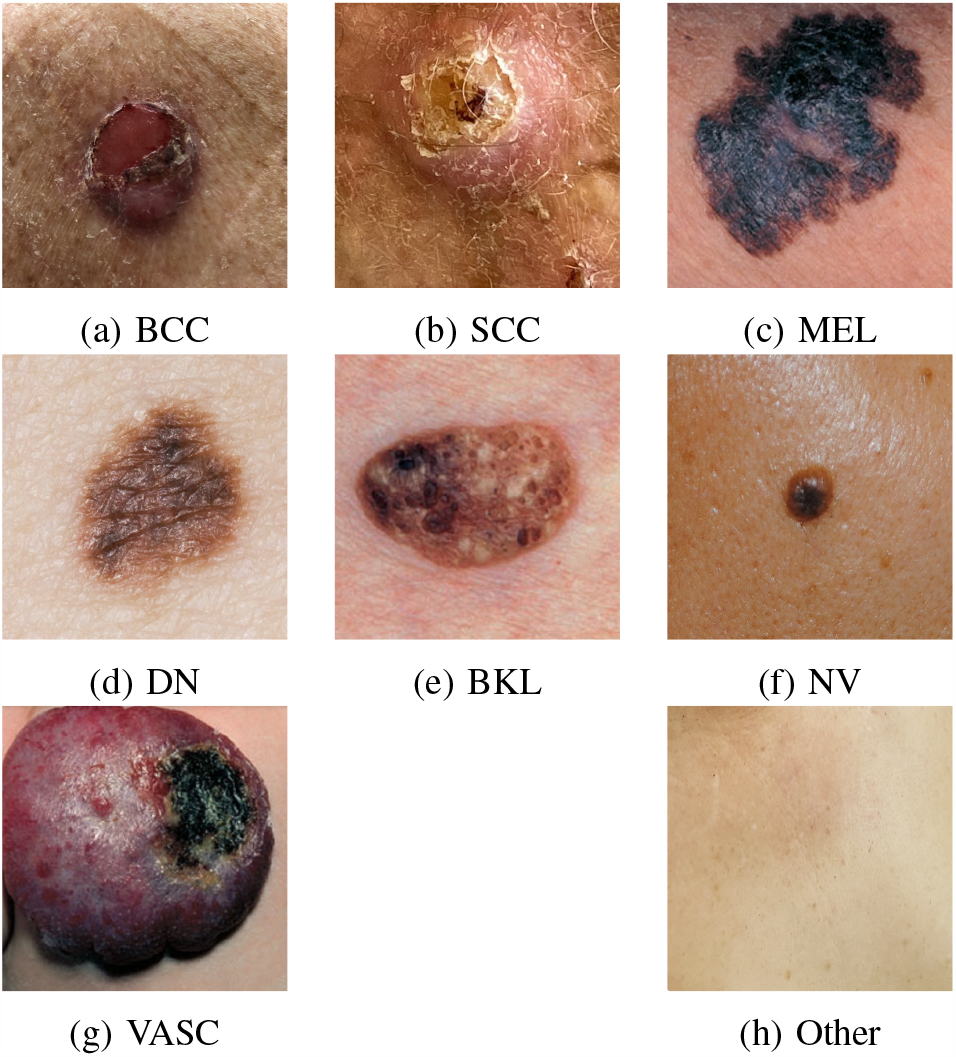
Examples of the skin lesion classes’ camera images.

We selected images corresponding to the considered classes from both datasets and merged them into one. We will refer to this dataset as dataset 3. Figure 2 depicts the distribution of the skin lesion classes in dataset 3.

**Fig. 2:**
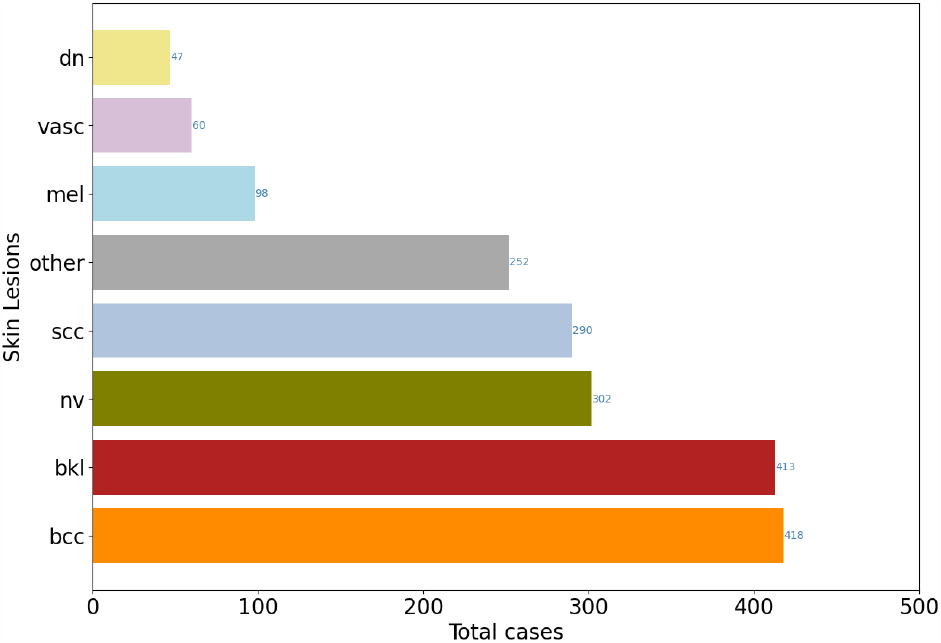
The distribution of the skin lesion classes in dataset 3.

It is important to mention that each of the images in dataset 3 was manually cropped, centered, and then resized to 224 by 224 pixels.

To further ensure the fairness of our models we conducted a statistical analysis of the existing skin tones in our dataset. We randomly sampled 320 images across all skin conditions (approximately 17% of the dataset). The images were then annotated using the Fitzpatrick skin types system [11]. Each image was labeled by a team of three human annotators. A single label was then assigned to each image based on the majority vote. Figures 3 and 4 depict the distribution of skin tones across all skin conditions and across cancerous skin conditions respectively. It can be clearly seen that lighter skin tones (Type 1, 2, and 3) are more prevalent than darker tones.

**Fig. 3:**
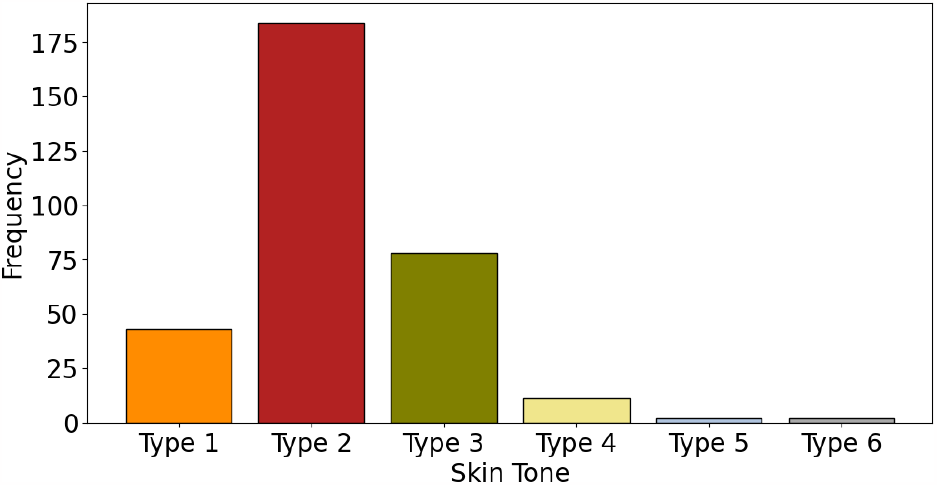
The distribution of the Fitzpatrick scale skin tones across all images.

**Fig. 4:**
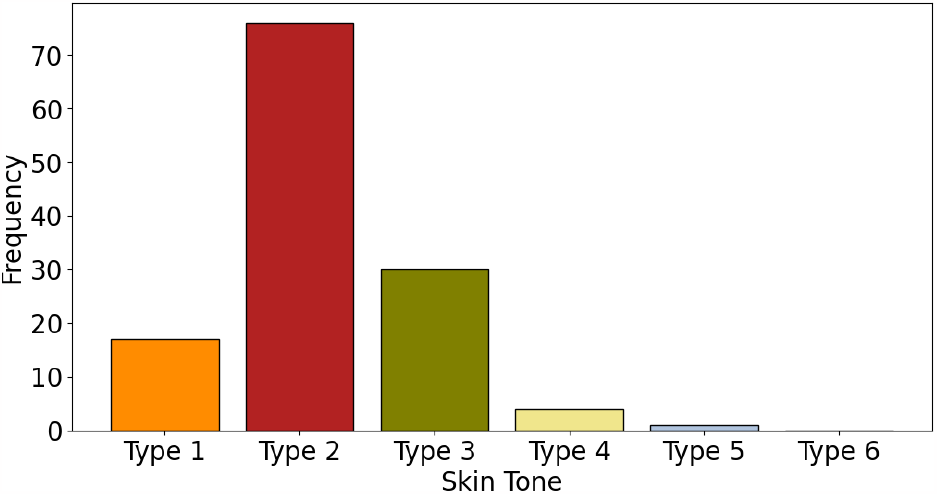
The distribution of the Fitzpatrick scale skin tones across cancerous skin lesion images.

### B. CNN architecture and transfer learning

We selected the Efficient Net architecture for the skin lesion classification model. Efficient Net models are a suite of CNN that outperformed many state-of-the-art CNN models such as Inception net, Resnet, Xception, etc [12]. Efficient Nets suite consists of seven models ranging from B0 to B7. These models were established by increasing their complexity in terms of depth, width, and resolution following a systematic scaling rule presented in formula 1.

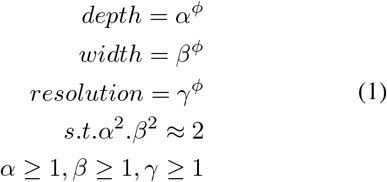

Where *α, β, γ* correspond to constants determined through grid search. While *ϕ* is the scaling factor.

Thus Efficient Net models were able to achieve optimal performance with minimal resources. In our project, we made use of Efficient Net B7 architecture. This model consists of seven blocks of neuronal layers. Each block contains various modules representing different combinations of convolution, pooling, and batch normalization layers.

We utilized the Efficient Net B7 model which was pretrained on the Canadian Institute For Advanced Research (CIFAR) dataset. We made modifications and added different types of layers to adjust it to our application. In the subsequent section, we give a thorough description of the training process.

### C. Training and Hyperparameter optimization

To train the skin lesion classification model, first, we split the preprocessed data into training and testing sets with an 80%:20% split ratio. 80% of the data was used for model training and the rest was holdout for model validation. We used stratified random sampling to randomly split the data into training and validation while conserving the original classes’ proportions. To further address the data imbalance issue, we assigned class weights that are inversely proportional to each skin lesion frequency. The larger class weights largely penalize the error on the less frequent classes which allows the model to better learn their patterns. CNN requires large amounts of data to learn meaningful patterns in images. Thus, we performed data augmentation on the training set to increase its size. This technique helps build robust models and combats overfitting in small datasets. The data augmentation consisted of applying different types of geometrical transformation to the images. These transformations include rotation, width and height shift, and horizontal and vertical flipping.

We conducted systematic optimization experiments to determine the most suitable architecture and set of hyperparameters. First, we dropped the last two layers of the pre-trained Effnet B7 model (the dropout and the dense layers). Next, we added various combinations of fully connected and dropout layers to adjust the Effnet model to our classification problem. We experimented with multiple combinations of values for the learning rate, the batch size, the number of units in the dense layer, and the dropout ratio. We also tested 224×224 and 600×0600 training images. The best architecture consisted of adding one dropout layer with a 0.2 dropout rate and one dense layer with the number of units corresponding to the number of classes. Lastly, we unfroze the last 111 layers of the pretrained Effnet B7 model. Overall the model has 29,942,376 trainable parameters and 34,175,799 Non-trainable parameters. We trained the model using the state-of-the-art ADAM optimizer. Figure 5 depicts an overview of the adopted Efficient Net B7 architecture.

**Fig. 5:**
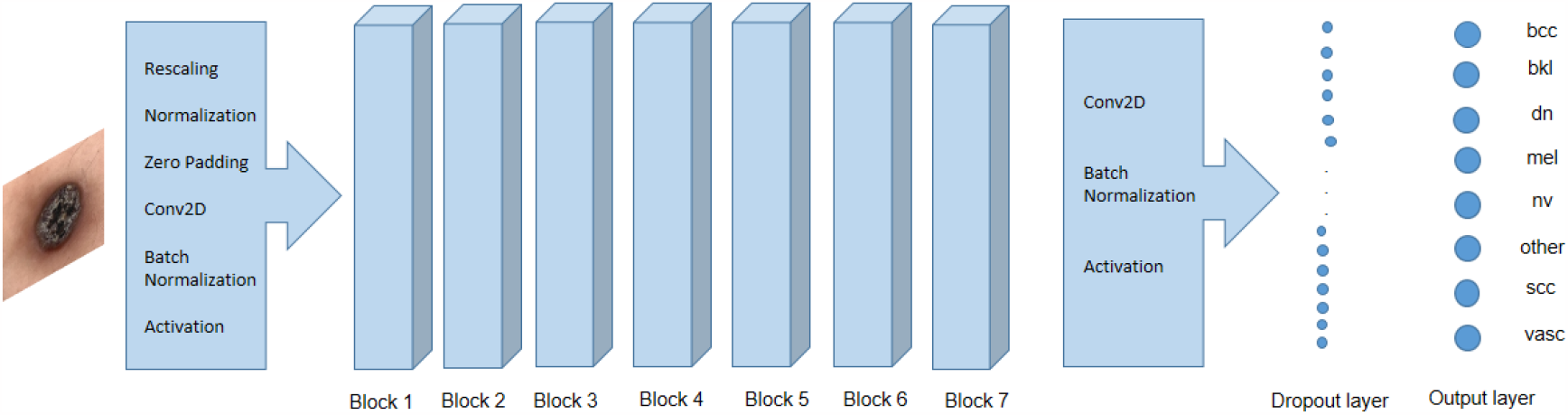
Efficient Net B7 architecture for skin lesion classification.

After conducting multiple training iterations the model performance was evaluated on the validation dataset and error analysis was performed to further understand and enhance the model’s predictive ability. The steps we followed are illustrated as a schema in Figure 6.

**Fig. 6:**
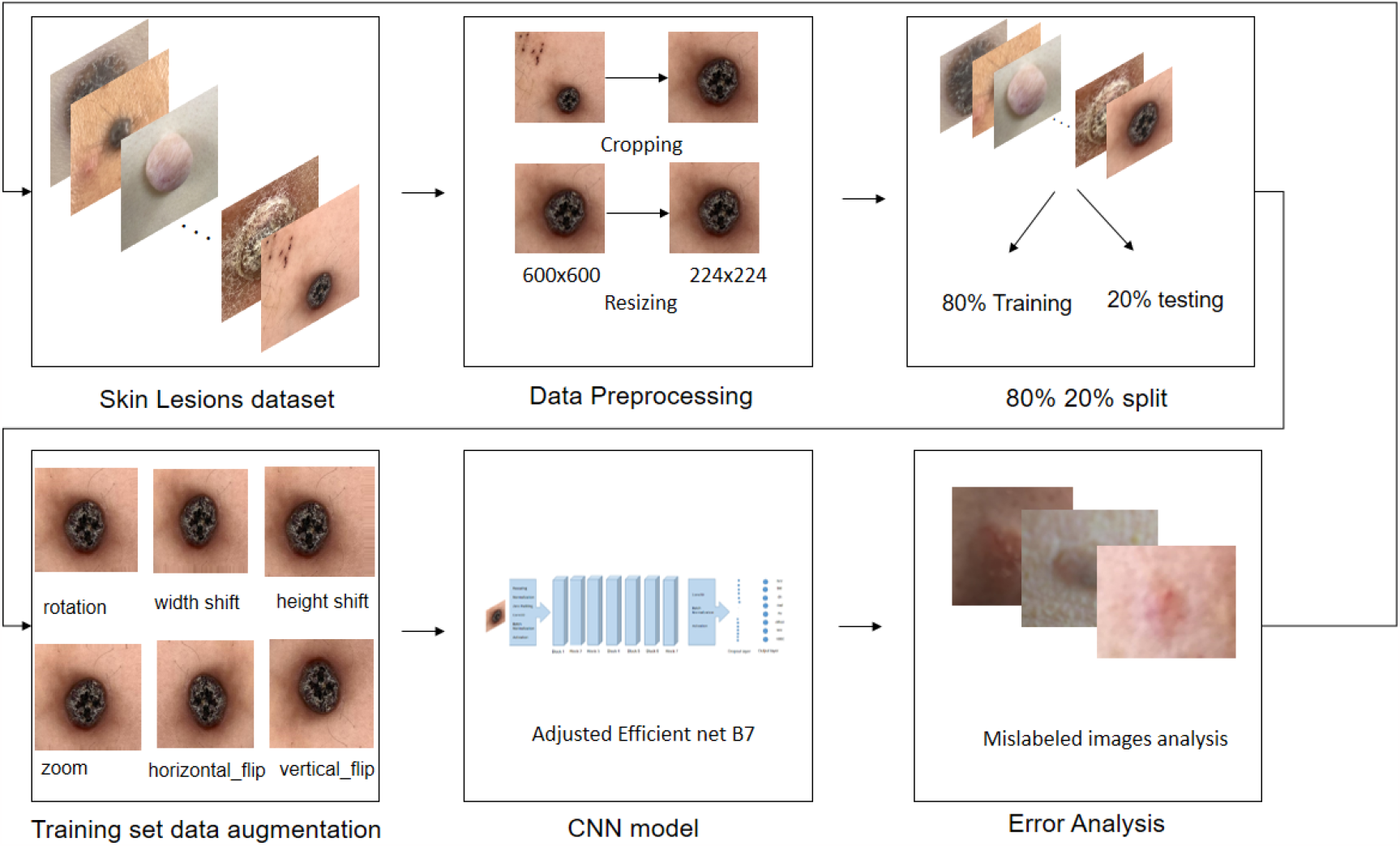
The Efficient Net model training and evaluation workflow.

### D. Implementation

For this project we used Python V 3.7.12 along with Google Colab Integrated Development Environment. For the data augmentation and the development of the CNN model, we used Tensor Flow 2.7 and Keras 2.7.

### E. Evaluation metrics

Several evaluation metrics were utilized to assess the performance of data-driven lesion classification models. The skin lesion data is characterized by a severe data imbalance as some of the skin conditions are less common than others. Due to this imbalance, almost all data-driven models demonstrate high performance in classifying frequent skin lesions and introduce the majority of errors in the less common conditions.

Thus, we considered three binary evaluation metrics: sensitivity, precision, and F1 score to assess the model’s performance on each skin condition separately. We have as well considered the overall model accuracy.

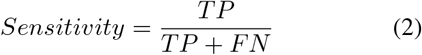

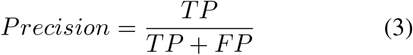

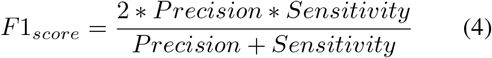

where:

- TP (True Positives): The number of the considered skin lesion images that were correctly identified.
- FP (False Positives): The number of the other classes’ images that were incorrectly predicted as the considered skin lesion.
- FN (False Negatives): The number of the considered skin lesion images that were incorrectly classified as the other skin lesions (non-detection error).

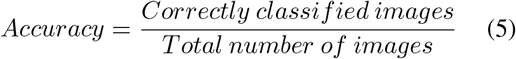

### F. Ad Hoc Testing

To understand the efficacy of the CNN-based classifier, we used patient-consented images (n=68; 36 malignant, 32 benign images; obtained with the Human Research Ethics Board approval: HREB.CC-20-0484) as a test dataset. These patients were clinically seen by 2 or 3 clinicians in Edmonton, AB before pathology determined the ground truth (malignant or benign). These patient images therefore have clinical evaluations and biopsies. We ran these images through our proposed multiclass classifier but categorized MEL, SCC, BCC, and DN as “malignant” lesions, and the rest of the lesions as “benign”, allowing us to do a binary classification analysis. The clinical and pathological analysis of these images were blinded for the classifier model-based analysis. Accuracy, specificity, and sensitivity were calculated, using three levels of stringencies, namely Top prediction and Top 3 predictions of the model, as well as averaging of the probability values of the 8 classes to give it a “malignant” or “benign” score. Clinical values were obtained by averaging the results from two or three human expert raters (dermatologists and plastic surgeons).

## III. Results and discussion

After multiple iterations of hyperparameter optimization, evaluation, and error analysis we found that 80% of the errors committed by the model correspond to blurry images. Figure 7 represents a sample of the mislabeled blurry images.

**Fig. 7:**
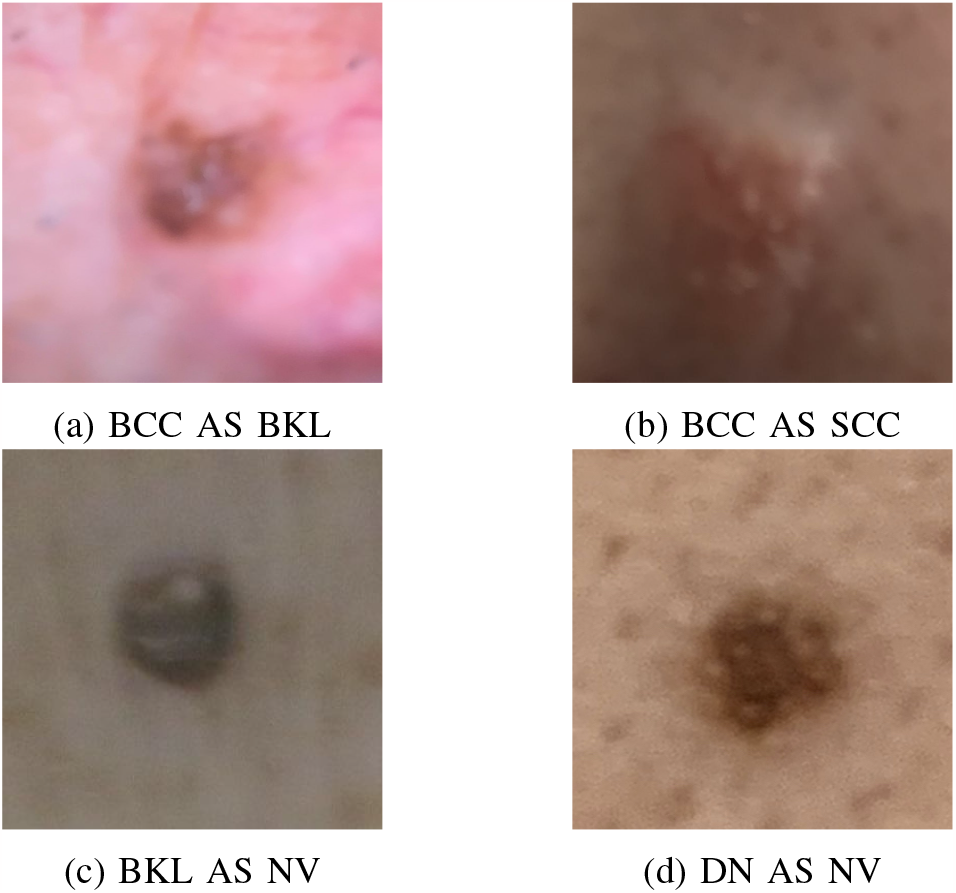
A sample of the mislabeled blurry images

After another iteration of data preprocessing including the removal of blurry images. The model achieved high classification performance with an accuracy of 87%, an average sensitivity of 83.75%, and an average specificity of 97.92%. Table I represents the precision, sensitivity, and F1 scores of the model on each of the predicted classes.

**TABLE I:**
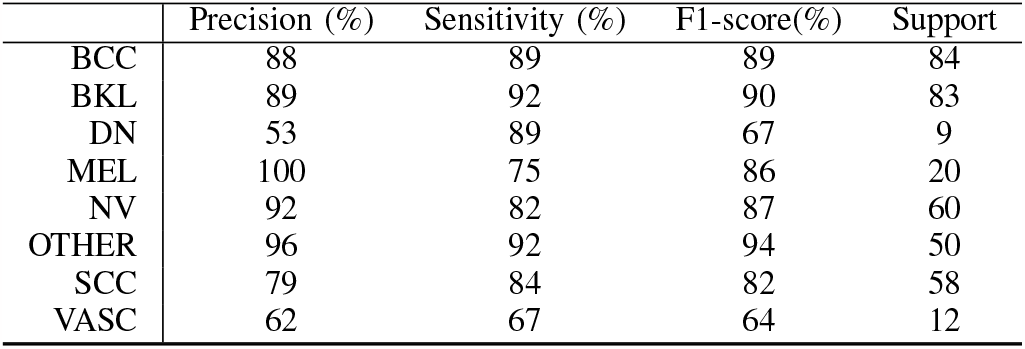
Precision, Sensitivity, and F1 scores of the skin lesion classification model on all skin conditions.

|  | Precision (%) | Sensitivity (%) | F1-score(%) | Support |
| --- | --- | --- | --- | --- |
| BCC | 88 | 89 | 89 | 84 |
| BKL | 89 | 92 | 90 | 83 |
| DN | 53 | 89 | 67 | 9 |
| MEL | 100 | 75 | 86 | 20 |
| NV | 92 | 82 | 87 | 60 |
| OTHER | 96 | 92 | 94 | 50 |
| SCC | 79 | 84 | 82 | 58 |
| VASC | 62 | 67 | 64 | 12 |

It can be clearly seen that the model achieves F1 scores higher than 80 for most of the classes except for DN and VASC. The low accuracy values in these classes are due to the small number of occurrences of these skin conditions compared to the others. To have a better understanding of the model’s performance figures 8 and 9 depict the Confusion matrix and the ROC curve.

**Fig. 8:**
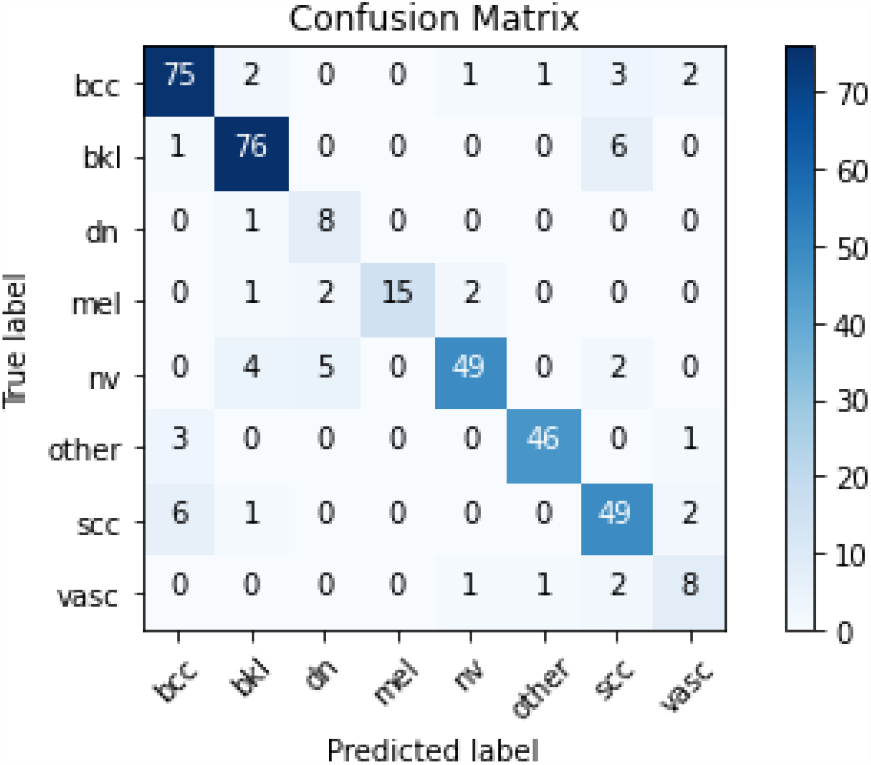
The Skin lesion model confusion matrix.

**Fig. 9:**
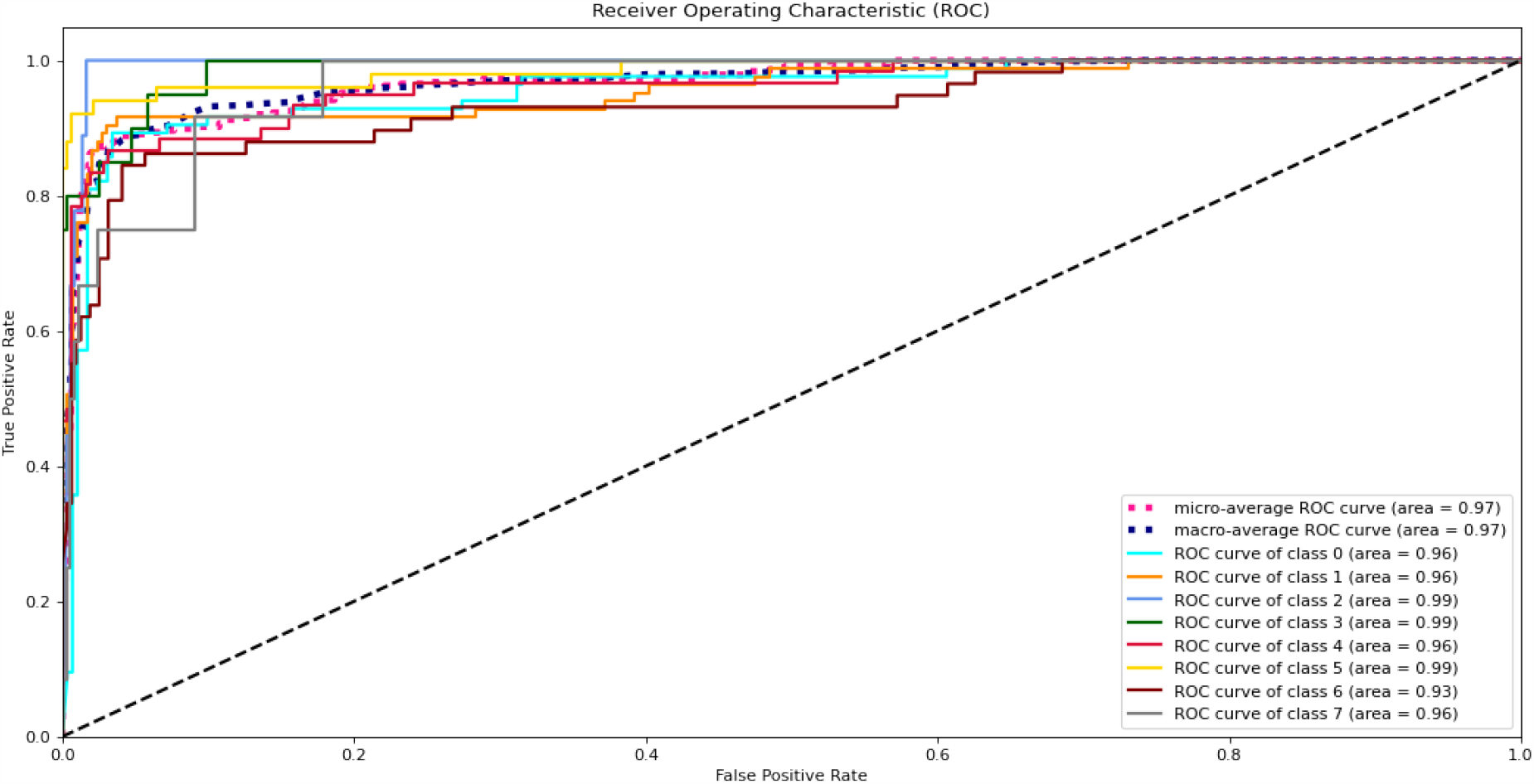
The ROC curves of the skin lesion classification model.

Based on Figure 8, we can see that the low precision in classifying DN was due to a high number of false positives. Specifically, MEL and NV were mislabeled as DN. This error is explainable by the big resemblance between these three conditions. However, as can be seen from the confusion matrix and confirmed by the ROC curve the model provides high classification performance. Acquiring more data and performing other training iterations will increase the model’s predictive performance and enhance its discriminatory power.

To further evaluate our proposed model, Table II shows the ad-hoc analysis results. it can be seen that the skin lesion classifier model attains an average accuracy of 78% versus a clinical average accuracy of 65%. The model sensitivity ranged from 75% to 88.9%, and specificity ranged from 65.3% to 81.25%. The average clinical sensitivity and specificity were 60% and 70%, respectively.

**TABLE II:**
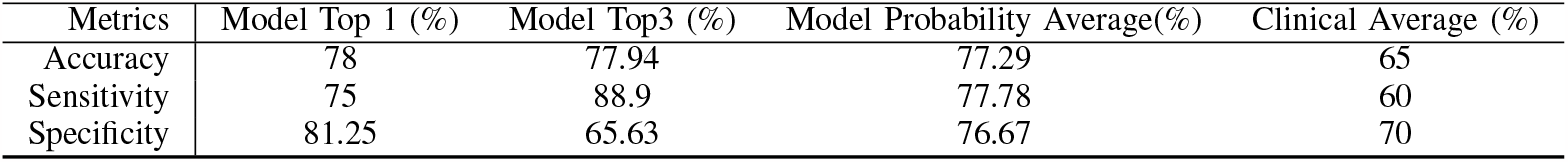
Accuracy, sensitivity, and specificity of the skin lesion classifier model and clinicians.

We observe the average accuracy of clinicians when doing a binary classification (malignant or benign) to be 65%. This falls within the range of accuracy of 54-85% for board-certified dermatologists, depending on clinical experience, as per the literature. Depending on the stringency of measure, the skin lesion classifier also outperformed the clinicians by 11%-28% (sensitivity) and 1%-11% (specificity). Although this retrospective ad hoc analysis, with small sample size and binary analysis, does not allow us to gain substantive statistical insights, including inter-rater comparisons, it does give us the preliminary data that shows the skin lesion classifier has clinical value, which can now be explored further by prospective efficacy studies.

## IV. Conclusion

Timely and early detection of skin cancer is essential to reduce patient morbidity and mortality rates. However, the literature review indicates that the clinical relevance of camera-based skin lesion image classification has not been fully addressed. In this study, we propose a skin lesion classification model based on Efficient Net B7 CNN. The model was trained and evaluated using a dataset consisting of images captured by a camera, which encompasses eight distinct classes of skin lesions: three cancerous types (BCC, SCC, and MEL) and five non-cancerous types (DN, BKL, NV, VASC, and Other). The experimental process included essential stages such as data preprocessing (involving cropping, resizing, and augmentation), optimization of model hyperparameters, and comprehensive error analysis. The outcome of this study revealed that the proposed model achieved a satisfactory classification accuracy of 87%. The unavailability of rare skin lesions and the quality of some camera images hinder the model performance from further improvement. To further enhance the model’s predictive ability, our future work includes acquiring more camera images, particularly those corresponding to rare skin lesions, with biopsy ground truths where available. Moreover, to ensure the fairness of our model, our future endeavors will include acquiring skin lesion camera images corresponding to darker skin tones.

## Data Availability

All data produced in the present study are available upon meeting patient consent guidelines

